# Estimating lung volumetric parameters via rapid, limited-slice, free-breathing thoracic dynamic MRI

**DOI:** 10.1101/2024.05.12.24306855

**Authors:** You Hao, Jayaram K. Udupa, Yubing Tong, Caiyun Wu, Joseph M. McDonough, Samantha Gogel, David M. Biko, Jason B. Anari, Drew A. Torigian, Patrick J. Cahill

## Abstract

**Purpose:** We present an observational study involving free-breathing short-scan-time dynamic MRI (dMRI) method that can be routinely used for computing dynamic lung volumes accurately.

**Materials and Methods:** (i) Full-resolution free-breathing sagittally-acquired 2D dMRI scans are gathered from 45 normal children via True-FISP sequence. Sparse dMRI (s-dMRI) scans are simulated from these datasets by subsampling in the spatio-temporal domains via a limited number *N_SS_* of selected sagittal locations and *T_SS_* of time instances (respectively, *N_FS_* and *T_FS_* for full scan). (ii) A 4D image is constructed from both full and sparse scans. Lungs are segmented from 4D image, and their volumes from full (*VF*) and sparse dMRI (*VS*) scans are computed. (iii) A regression model is fit for *VF* as a function of *VS* on a training set, and the full-resolution volume *VP* predicted by the model is estimated from *VS*. (iv) The deviation of *VP* from *VF* is analyzed on both synthesized sparse dMRI scans from a separate full-resolution test set and actual s-dMRI scans prospectively acquired from 10 normal children.

**Results:** With *N_SS_*=5 (per lung) and *T_SS_*=40, the deviation of *VP* from *VF* was ∼2% with a total scan-time of ∼9 min (45-60 min for the full scan with *N_FS_*=15-22 (per lung) and *T_FS_*=80). These metrics become 0.4%, and <20 min for s-dMRI with *N_SS_*=15-22 (per lung) and *T_SS_*=40.

**Conclusion:** s-dMRI is a practical approach for computing dynamic lung volumes that can be used routinely with no radiation concern, especially on patients who cannot tolerate long scan times.

## 1. Introduction

Assessment of dynamic volumes of the two lungs separately during tidal breathing is useful for assessment of many pediatric and adult thoracic disorders [1] such as scoliosis, thoracic insufficiency syndrome (TIS) [2, 3], and various pulmonary disorders [4], and for determining the effects of therapeutic interventions [5]. Several imaging modalities have been used for lung volume estimation, including chest radiography (CXR), thoracic computed tomography (CT), and thoracic MRI.

Lung volumes derived from CXR are based on the volumes contained within the thoracic contours [6–10]. The major limitation of this approach is its compromised accuracy stemming from estimating a 3D phenomenon via 2D projection spaces and the inability to properly separate left and right lungs. These issues become compounded for patients with deformed thorax and dynamic imaging.

Lung volumes derived from thoracic CT scans include tissue volumes and gas volumes [6] within the segmented region. Comparable correlations have been observed between chest CT as compared with plethysmographic total lung capacity [11–14]. However, patients would have to be able to perform an adequate breath-hold, which can be challenging for children. Alternatively, they would have to undergo image acquisition during general anesthesia to achieve breath holding, which is particularly problematic in the pediatric setting.

4D dynamic magnetic resonance imaging (dMRI) seems to be the best choice currently [15] because of its soft tissue contrast, lack of ionizing radiation, and greater flexibility in the choice of imaging plane for acquisition. However, its major limitation is that it requires a relatively long acquisition time [5, 15, 16], as it can take up to 45 minutes to complete a dMRI study. Therefore, approaches to acquire the images more rapidly without sacrificing the accuracy of volumetric parameters estimation are highly desirable.

We present a novel approach, called *sparse dMRI*, or *s-dMRI*, to significantly speed up a previously established dMRI scanning method for studying pediatric TIS [5, 15–22]. A very preliminary version of this work was presented in the 2021 SPIE Medical Imaging conference and its proceedings [23]. The present paper is substantially different with the following enhancements. (i) A detailed description of the methodology. (ii) Several strategies for both spatial and temporal sampling compared to just a basic method for spatial sampling. (iii) Extensive evaluation on retrospective and prospectively acquired data sets compared to demonstration on just a small retrospective data set. (iv) A comparative assessment with several related techniques.

## 2. Methods

### 2.1 dMRI scan protocol, subjects, image data

#### Full resolution scan

For each sagittal slice location through the chest, image slices are acquired rapidly while the subject breaths freely, using the following imaging parameters: 3T-MRI scanner (Verio, Siemens, Erlangen, Germany), True-FISP sequence, TR=3.82 ms, TE=1.91 ms, voxel size ∼1×1×6 mm^3^, 320×320 matrix, bandwidth 258 Hz, and flip angle 76 degrees, FOV: 15mm above apexes of lungs to base of kidneys. For each sagittal location, 80 slices are obtained over 8-14 breathing cycles at ∼480 ms/slice. Around 35 sagittal locations across the chest are imaged resulting in 2800-3200 2D slices. Total scan time is typically ∼45 min and up to an hour for larger subjects. From this set of slices, one representative 4D image comprising one full breathing cycle is constructed post hoc [15, 16] from which volumetric parameters are derived to demonstrate the utility of dMRI in studying TIS [5, 20–22]. The full resolution scan data were acquired from 45 normal pediatric subjects, including 26 females and 19 males (average age 9.5 years, range 6.2-13.7). These 45 subjects overlap with 20 subjects in the preliminary paper [23].

#### Sparse scan

In the sparse protocol, 10 subjects (4 males and 6 females with average age 16.3 years, range 13.1-18.8) were scanned using the above sequence, with 5-7 sagittal slices uniformly located across each of right lung and left lung and 40 temporal slices for each sagittal location. These 10 subjects were also scanned at full resolution for comparison.

All datasets were acquired under an ongoing research protocol approved by the Institutional Review Board at the Children’s Hospital of Philadelphia (CHOP) and University of Pennsylvania, along with Health Insurance Portability and Accountability Act authorization.

### 2.2 Overview of the framework

The s-dMRI approach is depicted in Figure 1. In the modeling stage, we utilize 20 training dMRI datasets from *full scans*, simulate spatio-temporal subsampling to create *sparse scans* from the full scans, compute various dynamic volumes, denoted *VF* from full scan and *VS* from sparse scan, and obtain a regression model to represent the relationship between *VF* and *VS*. In the prediction stage, we first compute *VS* from an independent cohort of sparse scan from 25 subjects, and then estimate the predicted full volume *VP* by using the prediction model. For details on the method, please see supplementary material.

**Figure 1.**
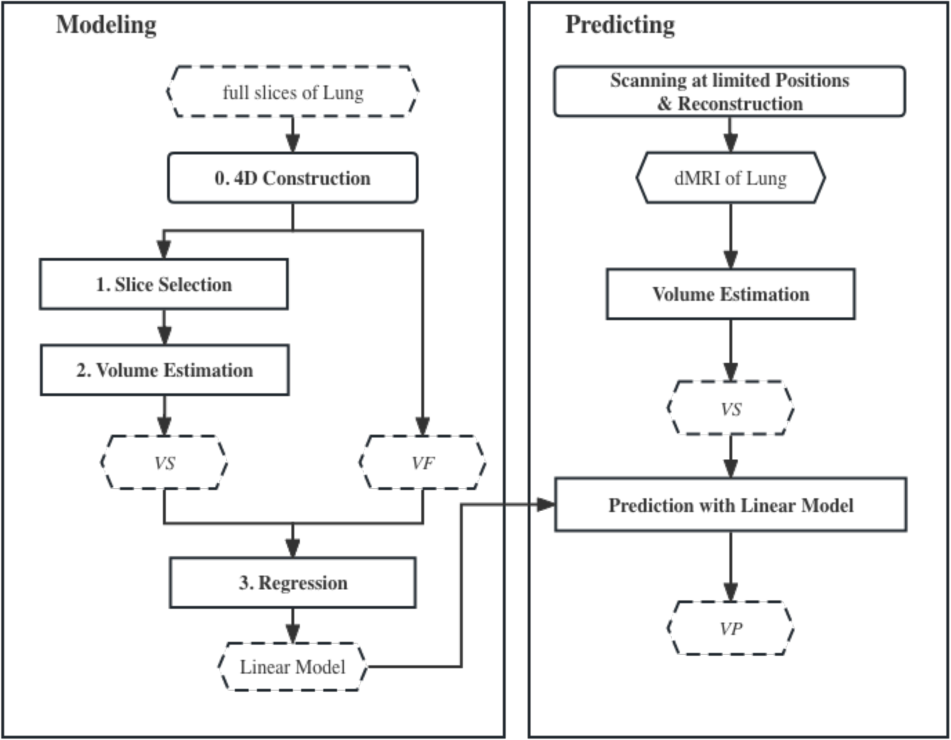
The framework of the proposed s-dMRI method to estimate lung volumetric parameters. In the modeling stage, a linear regression model is created between volume *VS* from sparse scans and volume *VF* from full scans using training data. In the prediction stage, *VF* is predicted (denoted by *VP*) from *VS* using the model and separate sparse scan data obtained either through simulation or via actual subject scans.

### 2.3 Spatial subsampling

There are three main parameters that determine spatial subsampling: <u>(i) Number of slices selected *N*</u>*<u>_SS_</u>*. The number of sagittal locations imaged in full scan, denoted *N_FS_*, varies from 15 to 22 slices between lateral and medial edges of each lung depending on body size. The idea of spatial subsampling is to select *N_SS_* < *N_FS_* locations between lateral and medial edges of each lung. <u>(ii) Method of selecting locations *MSL*.</u> We compared two strategies for selecting locations. In UFM strategy, *N_SS_* locations uniformly situated between the edges of the lung are selected for imaging. In MAX method, the sagittal location showing maximum lung cross-sectional area is first selected for each lung and subsequently the remaining locations are determined to be uniformly situated on the two sides of this location (Figure 2). Since UFM has no requirement of finding the location with the largest area, implementation of the method for actual clinical scanning is simple, practical, and straightforward. <u>(iii) Method of interpolation *Int*.</u> A straightforward way of determining lung volume *VS* from the sparse scan is via nearest neighbor (NN) interpolation. For a better approximation of the full scan volume *VF*, we compared three methods for interpolating the area curve to estimate *VS* – linear spline (LIN), quadratic spline (QUA), and cubic spline (CUB) (Figure 3).

**Figure 2.**
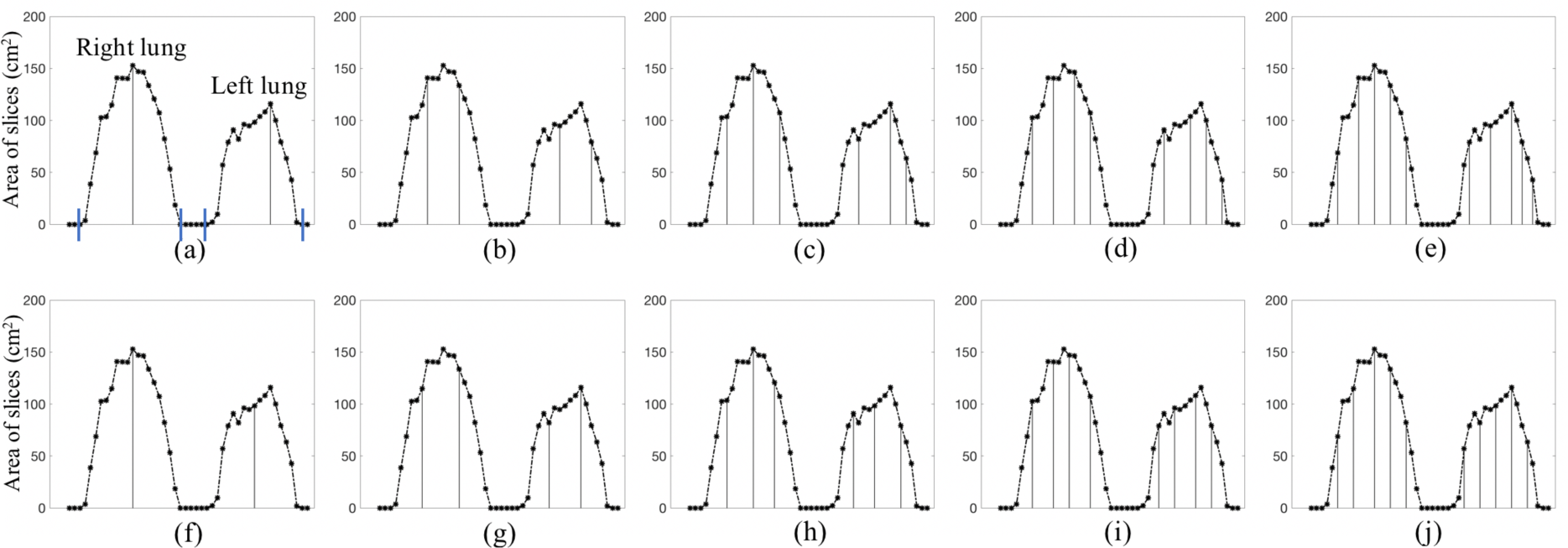
Illustration of the area curves for the two slice selection methods *MSL* = MAX and *MSL* = UFM. Sagittal locations are marked along the x-axis, starting from the right side of the patient with the first sagittal location slightly outside the thorax and ending with the last slice outside the left side of the thorax. The medial and lateral edges of the right and left lungs are marked in (a). (a-e) MAX: The numbers of selected slices *N_SS_* are respectively from 1 to 5. (f-j) UFM: The numbers of selected slices *N_SS_* are respectively from 1 to 5. The continuous curves represent the lung area as a function of the slice location obtained from the slices in the full scan. The vertical bars denote the area of the lung in the slice selected at that location in the sparse scan. *N_SS_* = Number of Slices Selected. *MSL* = Method of Selecting Locations. MAX = Maximum area method for *MSL*. UFM = Uniform method for *MSL*.

**Figure 3.**
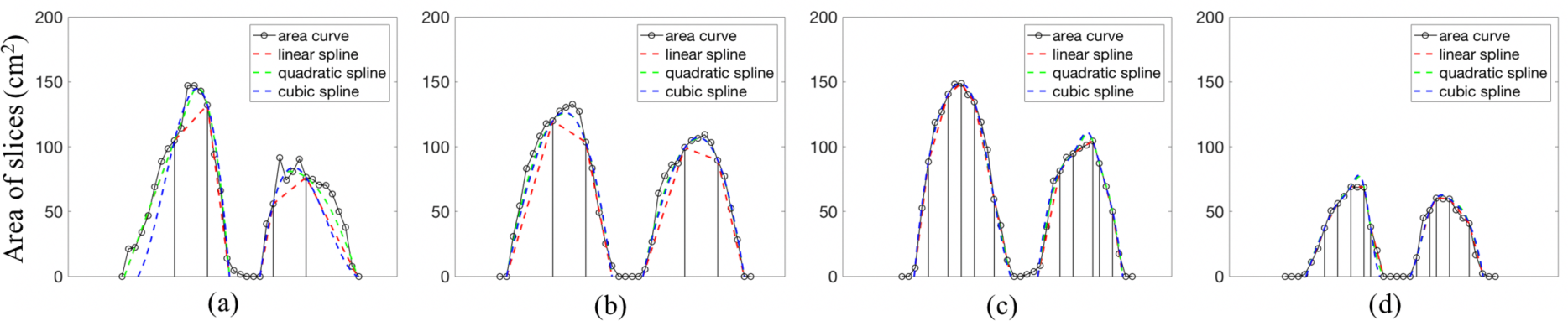
Illustration of the different interpolation methods on data sets from 4 different subjects. The layout is as in Figure 2. All examples use the MAX method of slice selection. Area curves shown: from full scan (black), LIN-linear spline (red), QUA-quadratic spline (green), and CUB-cubic spline (blue). In (a) and (b), *N_SS_* = 2; and in (c) an_d (d),_ *N_SS_ = 5. N_SS_* = Number of Slices Selected.

To build the regression model, we utilized the full-scan dMRI data and created sparse scans from them by selecting the sagittal locations. We designate the predicted lung volume derived from *VS* with the linear model as *VP* and use relative root mean squared (*rRMS*) error to evaluate the accuracy of the predictive model:

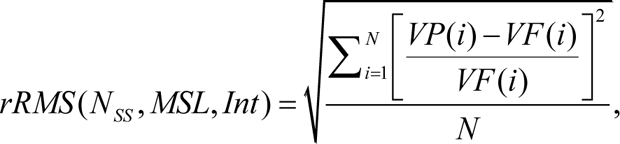

where *N* represents the number of samples used for the regression process.

Figure 4 illustrates the process of regression using UFM method with different *N_SS_* and interpolation methods. Notably, refined interpolation methods shown in Row 2 achieve better regression accuracy than NN shown in Row 1. Clearly, accuracy improves as *N_SS_* increases.

**Figure 4.**
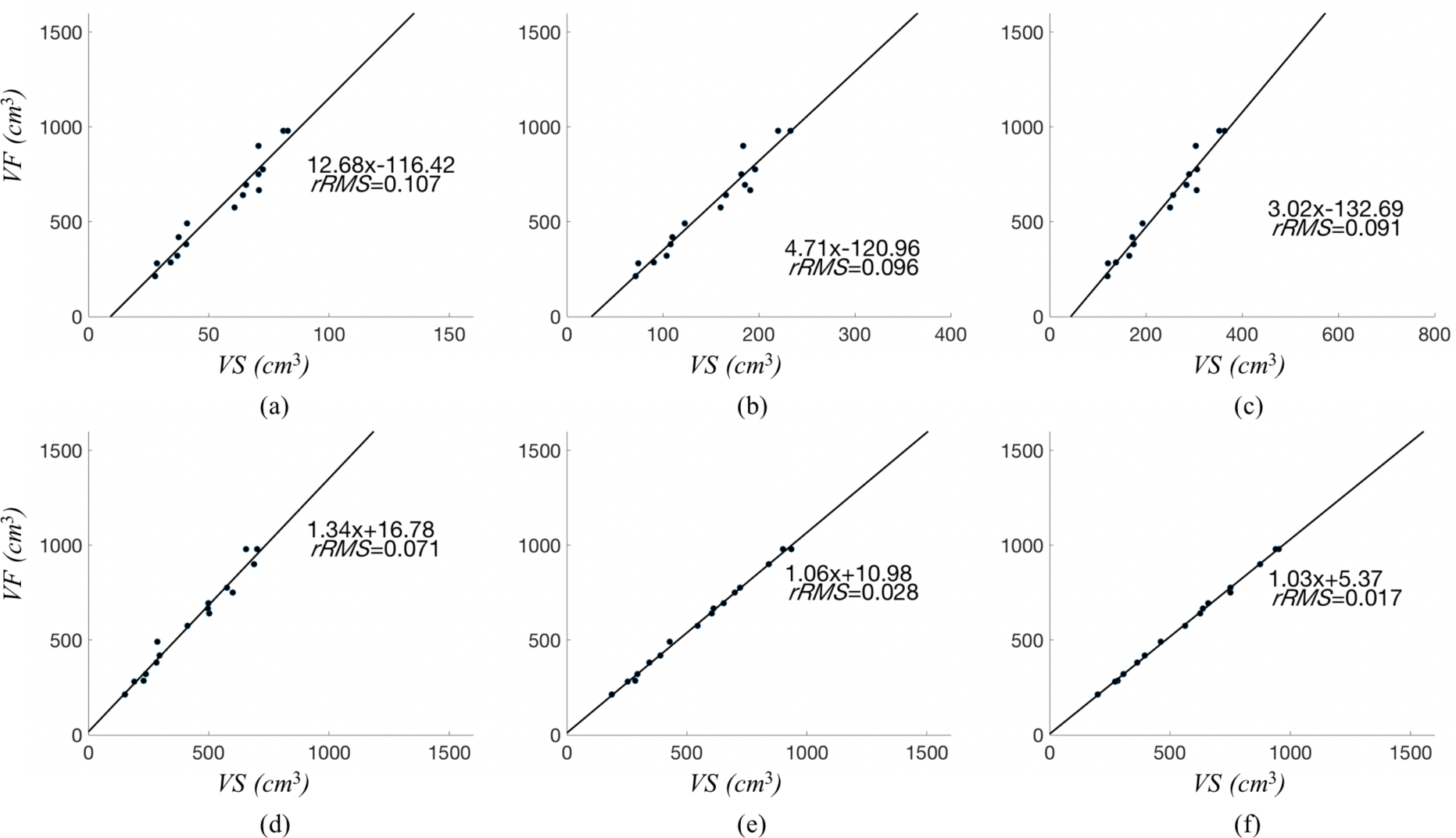
Illustration of linear regression from *VS* to *VF*. 20 samples of the left lung at end expiration from 20 normal subjects were used. The slice selection method used was *MSL* = UFM. 1st row: *Int* = NN. 2nd row: *Int* = LIN. (a), (d) *_NSS_* = 1; (_b), (e)_ *N_SS_* = 3; (c), (f) *N_SS_* = 5. *VF* = Volume from Full scan. *VS* = Volume from Sparse scan. *N_SS_* = Number of Slices Selected. *MSL* = Method of Selecting Locations. UFM = Uniform method for *MSL*.

### 2.4 Temporal subsampling

We denote the number of temporal slices acquired for each sagittal location in full scan by *T_FS_* (= 80 in in our case). We empirically evaluated the effect of choosing *T_SS_* < *T_FS_* time samples for the sparse scan on the changing lung volumes. In the full scan data with *T_FS_* time samples, we selected only the first *T_SS_* samples, perform 4D construction using these temporally “subsampled” data for different choices of *T_SS_*, and determined the volume deviation from the full scan as a function of the degree of temporal subsampling.

## 3. Results

### 3.1 Optimal parameter setting

The key independent measures we assess are right and left lung volumes at any time point such as end inspiration and end expiration, denoted, respectively, by RLVei, LLVei, RLVee, and LLVee. There are 4 parameters in the s-dMRI approach: *N_SS_*, slice selection methods (UFM or MAX), interpolation methods (NN, LIN, QUA, or CUB), and *T_SS_*. Of these, *N_SS_* and *T_SS_* behave similarly – the larger the value, the more accurate will be the estimated volumes. They are also the arbiters of the time savings compared to the full scan. Slice selection methods and interpolation methods determine the accuracy of approximation achieved for fixed values of *N_SS_* and *T_SS_*. Therefore, for different possible choices of *N_SS_* and *T_SS_*, we systematically study the tradeoff between accuracy and efficiency to choose the best setting for the parameters.

Figure S1 illustrates the variation of *rRMS* in spatial subsampling as a function of *N_SS_* with different slice selection methods and interpolation methods on the 20 data sets used for modeling for the four volumes RLVei, LLVei, RLVee, and LLVee. Since NN is non-competitive, it is excluded from consideration. Notably, for *N_SS_* ≥ 5, *rRMS* hovers around or falls below 2%, with LIN having a slight advantage over other interpolation methods. We may also note that there is not much difference in *rRMS* errors between slice selection methods MAX and UFM for *N_SS_* ≥ 5. Since UFM is simpler to actually implement in performing sparse scans, we recommend UFM as the method of slice selection.

Since *N_SS_* = 5 already achieves very low error, the changes we observe in volumes from pre- to post-operative condition in TIS treatment are typically greater than 20% [7], and increasing *N_SS_* beyond 5 lowers error insignificantly and gradually, for spatial subsampling we recommend the choice of *N_SS_* = 5, *MSL* = UFM, and *Int =* LIN, at least for the TIS application.

Figure S2 illustrates the variation in *rRMS* in temporal subsampling as a function of *T_SS_* on the 20 data sets used for modeling. Table 1 summarizes the actual *rRMS* values. Recall that for each value of *T_SS_*, full spatial volume is considered without any spatial subsampling to study the effect of temporal sub-sampling on its own. Notably, increasing *T_SS_* beyond 40 does not improve accuracy substantially and lowering the value below 40 is not advisable either. As such, we recommend *T_SS_* = 40 as the optimal setting.

**Table 1:** *rRMS* errors in volume estimation for different choices of the number of temporal samples *T_SS_* and no spatial subsampling.

| $T_{SS}$ | 20 | 30 | 40 | 50 | 60 | 70 |
| --- | --- | --- | --- | --- | --- | --- |
| LLVee | 0.020 | 0.017 | 0.015 | 0.011 | 0.009 | 0.008 |
| RLVee | 0.016 | 0.014 | 0.012 | 0.007 | 0.005 | 0.004 |
| LLVei | 0.028 | 0.020 | 0.013 | 0.014 | 0.011 | 0.008 |
| RLVei | 0.024 | 0.016 | 0.011 | 0.015 | 0.011 | 0.008 |

$r_{RMS}$ : relative Root Mean Squared error. LLVee: Left Lung Volume at End-Expiration. RLVee: Right Lung Volume at End-Expiration. LLVei: Left Lung Volume at End-Inspiration. RLVei: Right Lung Volume at End-Inspiration.

In the actual analysis of the accuracy of the whole s-dMRI process presented below, we will use the optimal settings of *MSL* = UFM, *Int =* LIN, and *T_SS_* = 40.

### 3.2 Assessing accuracy of the s-dMRI process

Using the optimal settings obtained as above, we determine the actual volume prediction accuracy in terms of *rRMS* error from retrospectively obtained data subsampled from full dMRI scans as well as prospectively acquired actual sparse scans.

#### Retrospective analysis

Figure 5 summarizes s-dMRI predictive performance in terms of *rRMS* error on the 25 test subjects for the 4 volume measures for the parameter setting of UFM, LIN, and *T_SS_* = 40 for different values of *N_SS_*. We also report the mean and standard deviations across the test subjects in Table S1. For this analysis, the sparse scans were obtained via simulation from the full scans. The mean scan time per lung is also shown in Figure 5 corresponding to different values of *N_SS_* as a dashed curve. The upper right corner of the curve represents the actual scan time per lung that was measured for *T_SS_* = 40 without leaving out sagittal locations and corresponding to roughly *N_SS_* = 15. In other words, the scan corresponding to that point involved temporal but not spatial sub-sampling. For lower values of *N_SS_*, the scan time shown is an estimation based on the number of slices imaged per lung. Notably, only with temporal sub-sampling (*T_SS_* = 40) and without any spatial sub-sampling, the total scan time can be reduced from ∼45 minutes to ∼20 minutes with an *rRMS* error of under 0.4% for the 4 key volume parameters compared to the full scan!

**Figure 5.**
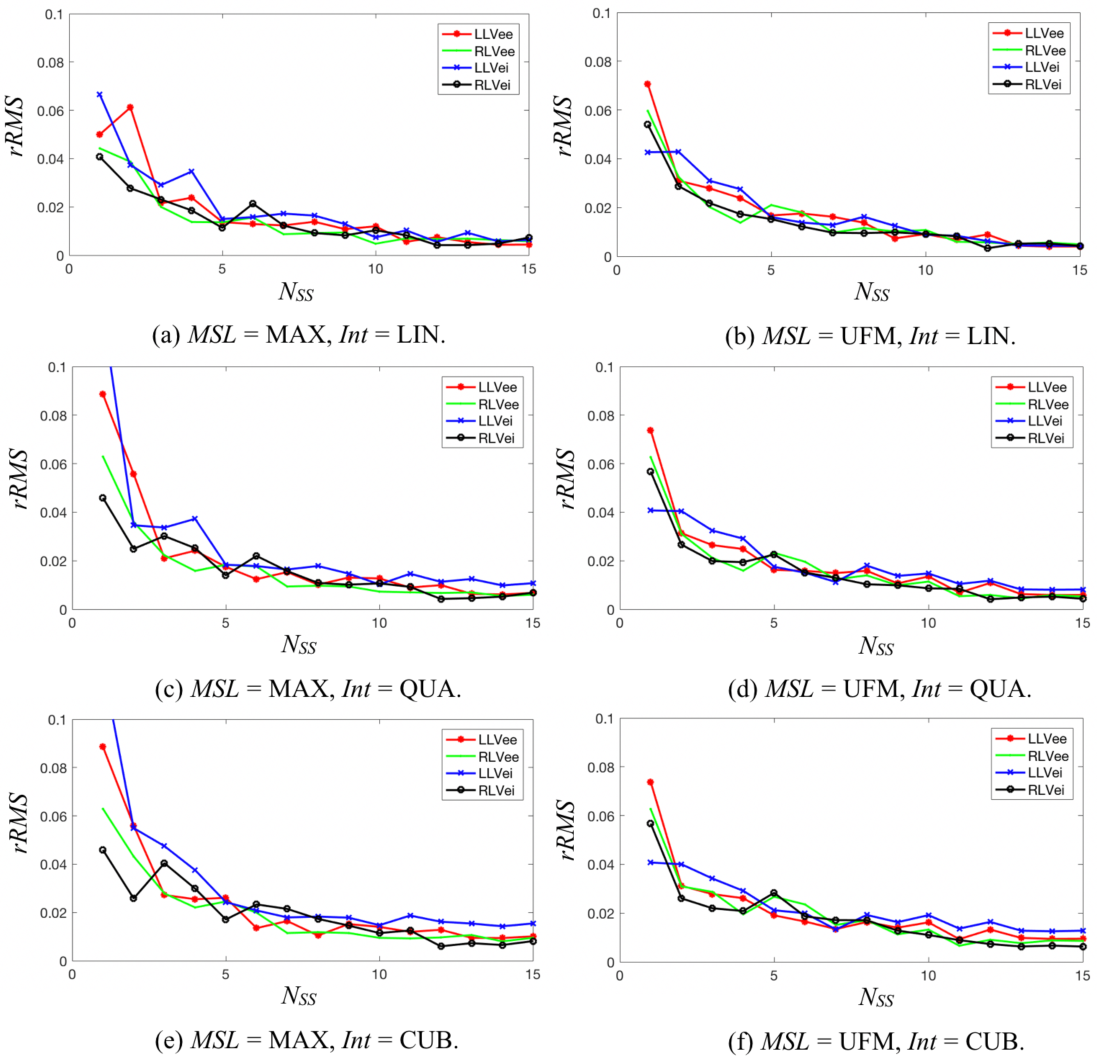
Illustration of relative root mean squared (*rRMS*) error of the s-dMRI process for estimating the 4 key lung volumes RLVei, LLVei, RLVee, and LLVee as a function of the parameter *N_SS_*. Errors are estimated from sparse scans simulated from full scans of 25 normal subjects. The curve of mean scan time per lung is also shown as a dashed curve. *N_SS_* = Number of Slices Selected. RLVei = Right Lung Volume at end-inspiration. RLVee = Right Lung Volume at end-expiration. LLVei = Left Lung Volume at end-inspiration. LLVee = Left Lung Volume at end-expiration.

#### Prospective analysis

For the prospectively acquired sparse scans (*N_SS_* = 5-7, *T_SS_* = 40), the *rRMS* errors in the different volume measures are as listed in Table 2. The reference measures used in this evaluation were the volumes obtained from the full-resolution scans of the same subjects. The mean scan time for the sparse scans was 4.5 min for each lung and the total scan time was ∼9 min. The error and the scan time are comparable to those from retrospective simulated scans for *N_SS_* = 5 and *T_SS_* = 40.

**Table 2.**
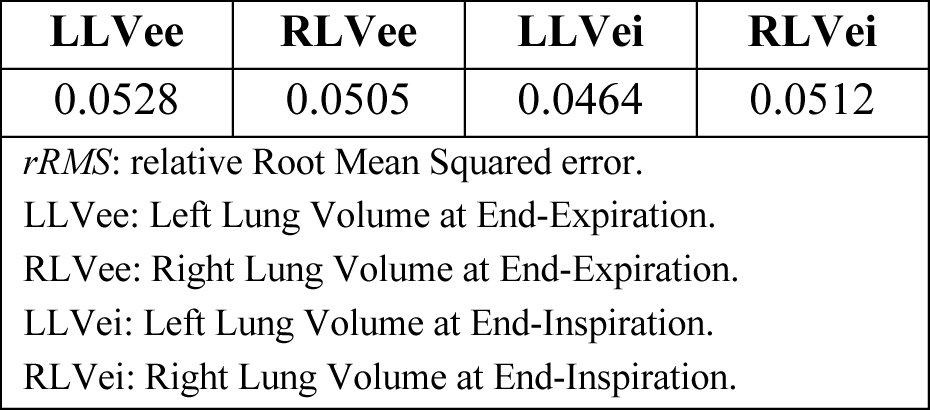
*rRMS* errors in volume measures in our prospective study involving sparse scans.

In Figure 6, we display the 2D spatial and temporal slices corresponding to the right lung selected from the 4D image constructed from a sparse scan of a normal female subject.

**Figure 6.**
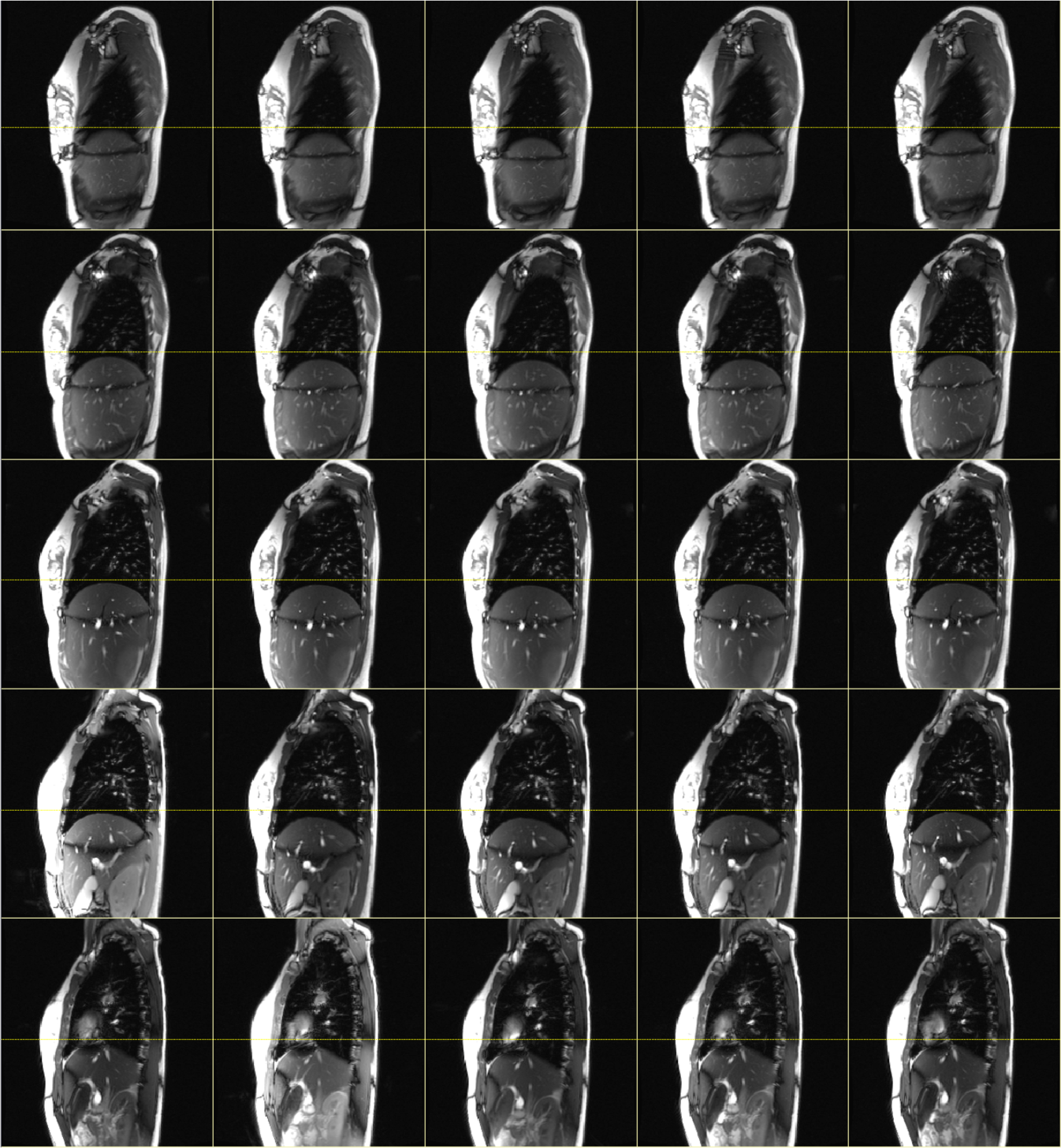
An example of the 4D constructed image of our sparse scan covering the right lung of a normal female subject. Each row shows one respiratory cycle of 5 time points for one sagittal location. The different rows indicate 5 different sagittal locations. The yellow dashed line represents the end-expiration level of the diaphragm dome.

## 4. Discussion

In this paper, we presented a novel approach using limited-slice, short scan-time, free-breathing, thoracic dynamic MRI to accurately estimate key lung dynamic volumetric parameters. Taking a well-established full resolution scan that takes 45 min – 1 hour per scan as reference, we demonstrated a root mean squared error in estimating volumes of less than 2% in pediatric subjects with a total scan time of ∼9 min by utilizing only 5 sagittal MRI slices through each lung and for each sagittal slice location acquiring 40 slices as a time sequence under free breathing conditions covering roughly 5-10 pediatric respiratory cycles. The method exploits the smoothness of the lung area in sagittal slices as a function of the slice location (Figures 2, 3) for each lung to achieve reduced scan time. By scanning at full spatial resolution (i.e., *N_SS_* =*N_FS_*) but reducing temporal samples by half (i.e., *T_SS_* = 0.5 *T_FS_*), we showed that the above error drops to ∼0.4% with a total scan time of ∼20 min. As such, the proposed s-dMRI approach constitutes a practical approach to measure dynamic volumes and their changes separately for each lung in any application that needs to study the dynamic function of the two lungs separately under natural free-breathing conditions. The approach can lead to increased patient comfort and convenience for practical real-world clinical applications, as we have observed in the TIS application. This may also potentially lead to improved image quality and usability due to a reduction of patient motion, discomfort, and abnormal breathing patterns

In the TIS application, the pre- to post-operative changes in the key volume parameters and tidal volumes are > 20% and > 40%, respectively [5], as such the above errors are unlikely to interfere with our ability to measure operative treatment response. Even the more accurate version with *N_SS_* = *N_FS_* and *T_SS_* = 40 is highly practical with a 20-minute scan time.

Dynamic MRI methods in the literature that demonstrated full 4D imaging capability under free-breathing conditions with demonstrated scan time practicality and accuracy metrics for measuring lung dynamic volumes in pediatric subjects are few. We summarize in Table 3 methods from the literature that are similar to the goals of this paper. Most existing techniques require breath maneuvering such as breath holding or separate navigator scanning or focus on analyzing the motion of the diaphragm only, often at a few selected points or slice locations, or for achieving adequate spatio-temporal resolution employ smaller field of view that does not cover the abdominal organs. Their conversion to estimate volumes routinely is not obvious or face additional challenges. In other words, a practical method for pediatric use without radiation concerns that can perform scanning in 10-20 minutes for estimating free-breathing dynamic volumes and tidal volumes separately for each lung does not seem to exist at present. Key competing methods are [24], which reports an ultra-short echo-time MRI, and XD-GRASP [25] and iMoCo [26] methods that provide adequate spatiotemporal resolution with about 5-10 minutes of acquisition time. However, the adequacy of image quality and coverage of the field of view in these methods for auto-segmentation of the lungs <u>and</u> key abdominal organs for pediatric applications remains to be proven. In our application, and hence in the full scan and sparse scan, the field of view extends to the inferior aspect of the kidneys since we are interested in studying how respiratory restrictions affect the mobility of key abdominal organs like the liver, kidneys, and spleen and vice versa, as already demonstrated [15, 16, 21, 22, 27, 28]. As such image quality and coverage should afford accurate auto-segmentation of not just the lungs but also these key organs. Studying the utility of such recent new imaging sequences for our purposes to reduce the acquisition time for each slice in our method but then use our current paradigm of weaving the selected slices together ingeniously through the OFx strategy [16] to form a 4D image will be one of our future goals.

**Table 3.** Summary of related papers from literature. NA = not available.

|  | Modality | Dimension | Breathing | Scan time | Population | # of subjects tested | Voxel size (mm<sup>3</sup>) |
| --- | --- | --- | --- | --- | --- | --- | --- |
| Qanadli et al., 1999 [29] | MRI | 2D | Breath Hold | 14s per slice | Adults (24-32) | 15 | 4.6x4.6 (mm<sup>2</sup>) |
| Coxson et al., 2008 [30] | CT | 3D | Breath Hold | Not available | Adults | 57 | 1.25x1.25x10 |
| Fuld et al., 2012 [31] | CT | 3D | Breath hold | < 10 s | Adults (20-64) | 37 | 0.61x0.61x0.5 |
| Jahani et al., 2015 [32] | CT | 4D | Breath hold | ~ 1 min | Adults | 6 | 0.5x0.5x0.75 |
| Higano et al., 2017 [24] | 3D radial UTE MRI | 3D | Free breathing | 9-16 min | Adults & neonates | 22 | 0.78x0.78x0.78 |
| Martini et al., 2019 [33] | dMRI | 2D | Free breathing | 2 resp cycles | Adults | 39 | NA |
| Safavi et al., 2020 [34] | MRI | 2D | Breath hold | 10 s per slice | Adults (>18 years) | 31 | NAxNAx6 |
| Sato et al., 2022 [35] | dMRI | 2D | Free breathing | 121 ms per image | Not available | 31 | 2x2x13.5 |
| XD-GRASP [25] | MRI | 3D | Free breathing | ~95s/partition x 80 partitions | Adult | 5 | 1.5x1.5x6 |
| iMoCo [26] | MRI | 3D | Free breathing | 5min30s | Adult Pediatric | 11 | 1.1x1.1x1.1 |

The main limitation of our study is the somewhat small sample size and the lack of demonstration of the s-dMRI approach in adults for broader applicability. Fortunately, since the respiratory rate is much lower in adults, we may be able to reduce *T_SS_* further to reduce acquisition time. However, it remains to be investigated if we may need to also increase *N_SS_* beyond 5, since the body width is greater in adults than in children, which may prolong the scan time.

In conclusion, s-dMRI offers a practical approach for computing dynamic volumes of individual lungs accurately that can be used routinely with no radiation concern, especially on patients who cannot tolerate long scan times. The s-dMRI sequence employed in this paper is a general sequence and can be easily implemented on any modern MRI scanner.

## Data Availability

All data produced in the present study are available upon reasonable request to the authors

## Supplemental Material

### S1. Details on Methods

#### S1.1 dMRI scan protocol, subjects, image data

There are two main approaches to form a 4D image through dMRI: (i) Real-time 3D volumetric approach: Use ultra-fast 3D MRI sequences to obtain real-time 3D volumetric data. (ii) Retrospective 2D slice approach: Use fast 2D MRI sequences to continuously acquire 2D images slice by slice while the subject is breathing freely, and then form a 4D image by selecting among these slices. For real-time methods, given the limitations of current hardware and software, it is difficult to achieve high spatial and contrast resolution, high signal-to-noise ratio, and sufficiently high temporal resolution simultaneously, especially given the high respiratory rate of pediatric subjects, which may compromise our ability to reliably perform auto-segmentations. Therefore, in this paper, for the s-dMRI method, we utilized the second approach for image acquisition.

In this approach, for each sagittal slice location through the chest, covering from the right edge of the thoracic body region to the left edge, image slices are acquired rapidly while the subject breaths freely and normally, using the following imaging parameters: 3T MRI scanner (Verio, Siemens, Erlangen, Germany), True-FISP sequence, TR=3.82 ms, TE=1.91 ms, voxel size ∼1×1×6 mm3, 320×320 matrix, bandwidth 258 Hz, and flip angle 76 degrees, field of view extending from at least 15 mm above the apex of the lungs superiorly to the base of the kidneys inferiorly. For each sagittal location, 80 image slices are obtained over 8-14 tidal breathing cycles at ∼480 ms/slice time-continuously. On average, 35-40 sagittal locations across the chest are imaged. Therefore, a total of 2800-3200 2D MRI slices are generated which constitute a *full-resolution* spatio-temporal sampling of the subject’s dynamic thorax over 100s of respiratory cycles. Total scan time is typically ∼45 min and up to an hour for larger subjects. From this set of slices, one representative 4D image comprising one full breathing cycle of the subject, constituted by 200-300 slices, is constructed post hoc via image analysis strategies [S1, S2].

In the study of TIS, to the 4D image constructed from the above full-resolution scan, we apply a sequence of operations to derive several volumetric parameters including left and right lung volume at end inspiration (EI) and end expiration (EE), and tidal volumes of the two lungs [S3-S7]. Our goal in the s-dMRI method is to maximally reduce the number of slices acquired both spatially and temporally so that the total scan time is significantly reduced without compromising the accuracy of estimating these dynamic volumes.

dMRI datasets for this paper are acquired from 45 normal pediatric subjects, including 26 females and 19 males (average age 9.5 years, range 6.2-13.7). Additionally, for assessing the change in volume estimation that may occur in the same subject from full resolution dMRI to s-dMRI, 10 subjects (4 male and 6 female) were scanned using the above full-resolution protocol <u>and</u> a sparse protocol where 5-7 sagittal slices uniformly located across each of the right lung and left lung were acquired with only 40 temporal slice data gathered for each sagittal location. All datasets were acquired under an ongoing research protocol approved by the Institutional Review Board at the Children’s Hospital of Philadelphia (CHOP) and University of Pennsylvania, along with Health Insurance Portability and Accountability Act authorization. Of the scans from 45 subjects, 20 cases were used for the “modeling” aspect of the s-dMRI approach, and the remaining 25 cases were used for predictive testing.

#### S1.2 Overview of the framework

The pipeline of our s-dMRI approach is depicted in Figure 1. In the modeling stage, we utilize the 20 training dMRI datasets from *full scans* through the thorax, simulate spatio-temporal subsampling to create *sparse scans* from the full scans, compute various dynamic volumes, denoted *VF* from full scan data and *VS* from sparse scan data, and obtain a regression model to represent the relationship between *VF* and *VS*. In the prediction stage, we first compute *VS* from an independent cohort of test sparse scan data from 25+10 subjects, obtained either through simulation or via actual subject scans, and then estimate the predicted full volume *VP* by using the created prediction model. Our aim is to make *VP* as close as possible to *VF.* The 4 key independent volumetric parameters needed in our application are: left and right lung end-inspiratory volumes and left and right lung end-expiratory volumes. We compute volumes *VF* and *VS* from full and sparse scans from the segmentations produced by a deep-learning-based method [S7], which achieved a mean ± standard deviation of Dice of 0.97±0.02, and performing manual correction as needed.

We think of the current scanning set up as *full scan* constituting full resolution acquisition since the acquisition time is already at the limit of practical viability and increasing the spatial and/or temporal resolution further will only add more time. There are two aspects to sparse scanning for reducing dMRI acquisition time: subsampling in the spatial domain – meaning selecting fewer sagittal locations and subsampling in the temporal domain – meaning acquiring less than 80 temporal slices for each sagittal location. The 4D image construction method we employ [S2] builds from these data (whether full or sparse scan) one 4D image representing the subject’s breathing thorax over one breathing cycle. This is accomplished via an optical-flux-driven optimization method. Firstly, the breathing signal for each sagittal location is extracted based on the flux of the optical flow vector field of the body region from the image time series. The optical flux allows us to perform a full analysis of all respiratory cycles, extract only normal cycles in a robust manner (and discard abnormal cycles such as those due to shallow or deep breathing), and map all extracted normal cycles on to one respiration model cycle for each sagittal location. The normal cycle models associated with the different sagittal locations are finally composited to form the final constructed 4D image (see [S2] for details). Lowering the number of spatial locations will result in poorer spatial representation of the lungs while lowering the number of time instances can lead to poorer representation of the dynamics in the 4D image. We will study these subsampling processes, their trade-off, and our volume estimation strategy in the following sections.

#### S1.3 Spatial subsampling

Our approach is to select fewer sagittal locations over the lung regions strategically for imaging and then fit a smooth function to represent the area of the lung at the selected slices as a function of the slice location so as to incur minimal error in volume estimation compared to volume from full resolution images. There are three main parameters that determine this selection: (i) the number of slices selected, *N_SS_*, (ii) the method of selecting these locations, *MSL*, and (iii) the interpolating function *Int* used to fit the area curve as a function of the slice locations. We assume that the sagittal locations corresponding to the lateral and medial edges of each lung are identified and known.

##### Number of slices selected *N_SS_*

The number of sagittal locations imaged in our current full scan, denoted *N_FS_*, varies from subject to subject, depending on the body size, from 15 to 22 slices between the lateral and medial edges of each lung. The idea of spatial subsampling is to select *N_SS_* < *N_FS_* locations between the lateral and medial edges of each lung. This parameter maximally influences accuracy. Accuracy also depends on the other two variables, as demonstrated in Figures 2 and 3.

##### Method of selecting locations *MSL*

We compared two strategies for selecting locations, named UFM and MAX. In the uniform strategy UFM, *N_SS_* locations uniformly situated between the lateral and medial edges of the lung are selected for imaging. In the MAX method, the sagittal location showing maximum lung cross-sectional area is first selected for each lung and subsequently the remaining locations are determined to be uniformly situated on the two sides of this location from the lateral or medial edge. See Figure 2. In our experiments, for the MAX method, we first find the sagittal locations that are at the lateral and medial edges of each lung in the localizer image. Then we find the sagittal location that roughly shows the largest area for the lung in the localizer image. The number of locations desired for the two segments in either side of this location is then specified. For the UFM method, the approach is similar without the requirement of finding the location with the largest area, but the total number of desired locations is specified. Thus, implementation of the method for actual clinical scanning is simple, practical, and straightforward.

##### Method of interpolation *Int*

A straightforward way of determining lung volume *VS* from the sparse scan is via nearest neighbor interpolation – compute volume contributed by each selected slice by multiplying the area of the lung in the slice by the space between slices and add up these volumes over all selected slices. This is equivalent to nearest neighbor (NN) interpolation. For a better approximation of the full scan volume *VF*, we compared three methods for interpolating the area curve to estimate *VS* – linear spline (LIN), quadratic spline (QUA), and cubic spline (CUB) (Figure 3). The idea is that the estimated volume is obtained by integrating the areas under the area curve which results from interpolation (see Figure 3). In Figures 3(a) and (b), *N_SS_* is too small, and LIN interpolation is not as good as the higher-order spline interpolation methods. In Figures 3(c) and 3(d), however, there is not much difference among the three interpolation methods. As the number of selected slices (*N_SS_*) increases, the accuracy of approximation will improve although the image acquisition time will also increase, as there is a trade-off between the accuracy of approximation and image acquisition time.

##### Regression

To build the regression model, we utilized the full-scan dMRI data we already had and created sparse scans from them by selecting the sagittal locations as explained above. This allowed us to perform various simulation experiments via different *N_SS_*, location selection methods, and interpolation methods without having to actually perform sparse scans on subjects. We utilized regression models built via simulated sparse scans in this manner and tested the s-dMRI method on both simulated scans that were not used for modeling and actual prospectively acquired sparse scans.

We designate the predicted lung volume derived from *VS* with the linear model as *VP* and use relative root mean squared (*rRMS*) error as a function of *N_SS_*, *MSL*, and *Int* to evaluate the accuracy of the predictive model:

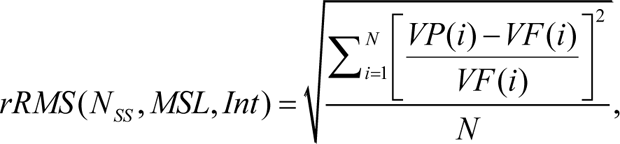

where *N* represents the number of samples used for the regression process.

Figure 4 illustrates the process of regression using UFM method and different *N_SS_* and interpolation methods. Notably, refined interpolation methods shown in Row 2 achieve better regression accuracy than NN shown in Row 1. Furthermore, accuracy improves as *N_SS_* increases, and even when *N_SS_* = 1 (Figures 4 (a), (d)), there is a strong linear relationship between *VS* and *VF*. Note how we exploited the smoothness of the area curve for each lung separately to reduce the number of slices and hence the acquisition time. The sagittal slice planes are ideal in this regard compared to axial or coronal.

#### S1.4 Temporal subsampling

We denote the number of temporal samples (slices) acquired for each sagittal location in our current full scan by *T_FS_*. As explained previously, *T_FS_* = 80 in the full resolution dMRI protocol. The idea behind temporal subsampling is to acquire *T_SS_* < *T_FS_* time samples so as to reduce acquisition time <u>and</u> incur minimal loss of accuracy in estimating lung volumes.

The temporal resolution achievable in our dMRI full scan, or equivalently the number of time instances captured over one respiratory cycle in our constructed 4D image, is determined by several factors: the respiratory rate (*RR*) of the individual subject, the time needed for acquiring one image slice data (480 ms in our current protocol), and the number of normal tidal breathing cycles observed over *T_FS_* time samples. Our 4D construction method OFx first identifies (slices constituting) the normal breathing cycles and compiles the distinct respiratory phases observed in these cycles into one composite respiratory cycle (see [S2] for details). The temporal resolution thus improves as we gather more normal cycles in a subject scan. When *RR* is high, the number of normal cycles collectable and hence the number of time instances captured in the 4D image becomes low. Conversely, when *RR* is low, more normal cycles are collected, resulting in higher temporal resolution in the 4D image. In our current full resolution scan, we achieve 4-10 time instances in the constructed 4D image, as observed in subjects of age 4 to 20 years.

Given the above theoretically intractable complex variabilities, we decided to perform an empirical evaluation of the effect of choosing *T_SS_* < *T_FS_* time samples on the changing lung volume. In the full scan data with *T_FS_* time samples we already have, we select only the first *T_SS_* < *T_FS_* samples, perform 4D construction using these temporally “subsampled” data for different choices of *T_SS_*, and determine the volume deviation from the full scan as a function of the degree of temporal subsampling.

#### S1.5 Evaluation strategies

##### Evaluation

Optimal setting for the model: The accuracy of volume estimation for a given time point is determined by the three spatial subsampling parameters *N_SS_*, slice selection methods, and interpolation methods. As such, we first fix the time point to EE and EI and analyze the model fit in terms of *rRMS* as we vary *N_SS_*. Similarly, for different choices of *T_SS_* we perform 4D construction for full scan spatial sampling and study the behavior of *rRMS* as a function of *T_SS_*. From these analyses, we determined the setting for the 4 parameters which would yield minimal *rRMS* error for the desired speed of dMRI scanning.

Assessing accuracy of the whole s-dMRI process: Using the optimal setting obtained as above, we determine the actual volume prediction accuracy in terms of *rRMS* error from retrospectively obtained data subsampled from full dMRI scans as well as prospectively acquired actual sparse scans.

### S2. Extra Tables and Figures

**Table S1**: Mean and Standard deviations of the relative errors.

**Figure S1**. Illustration of relative root mean squared (*rRMS*) error of model fit for the 4 static lung volumes RLVei, LLVei, RLVee, and LLVee as a function of the parameters *NSS*, *MSL*, and *Int* in the modeling stage.

**Figure S2**. Illustration of relative root mean squared (*rRMS*) error of model fit for the 4 volumes RLVei, LLVei, RLVee, and LLVee and as a function of the parameter *TSS* in the modeling stage.

---

**Table S1:** Mean and Standard deviation of relative root mean squared (*rRMS*) errors for different choices of number of slices in the sparse scan *N_SS_*. Here method of selecting locations *MSL* = UFM (uniform) and method of interpolation *Int* = LIN.

| $N_{ss}$ | **LLVee** | **RLVee** | **LLVei** | **RLVei** |
| --- | --- | --- | --- | --- |
| **1** | 0.0418±0.0392 | 0.0544±0.0387 | 0.0453±0.0483 | 0.0475±0.0428 |
| **2** | 0.0341±0.0258 | 0.0221±0.0193 | 0.0249±0.0175 | 0.0302±0.0227 |
| **3** | 0.025±0.0204 | 0.0237±0.0168 | 0.0269±0.0178 | 0.0179±0.014 |
| **4** | 0.0193±0.0124 | 0.0208±0.0127 | 0.0209±0.0114 | 0.0149±0.0086 |
| **5** | 0.0182±0.0124 | 0.0153±0.0078 | 0.0162±0.0127 | 0.0174±0.0124 |
| **6** | 0.0184±0.0093 | 0.0129±0.0091 | 0.0132±0.0076 | 0.0134±0.0098 |
| **7** | 0.0137±0.0102 | 0.0112±0.008 | 0.0132±0.0091 | 0.0117±0.0088 |
| **8** | 0.0187±0.0133 | 0.011±0.0088 | 0.0113±0.0079 | 0.0142±0.0098 |
| **9** | 0.0144±0.0085 | 0.0098±0.0093 | 0.0091±0.0072 | 0.0134±0.0089 |
| **10** | 0.0118±0.0102 | 0.0086±0.0067 | 0.0096±0.0095 | 0.0103±0.0079 |
| **11** | 0.0111±0.0096 | 0.0095±0.0063 | 0.0098±0.0092 | 0.0118±0.009 |
| **12** | 0.0124±0.0096 | 0.0082±0.007 | 0.0097±0.0082 | 0.0108±0.0082 |
| **13** | 0.0113±0.0101 | 0.0085±0.008 | 0.0102±0.0073 | 0.0098±0.008 |
| **14** | 0.0112±0.0097 | 0.0081±0.0073 | 0.0094±0.0075 | 0.0094±0.0076 |
| **15** | 0.0111±0.0096 | 0.0081±0.0072 | 0.0092±0.0074 | 0.0091±0.0078 |
| LLVee: Left Lung Volume at End-Expiration.<br>RLVee: Right Lung Volume at End-Expiration.<br>LLVei: Left Lung Volume at End-Inspiration.<br>RLVei: Right Lung Volume at End-Inspiration. |  |  |  |  |

**Figure S1.**
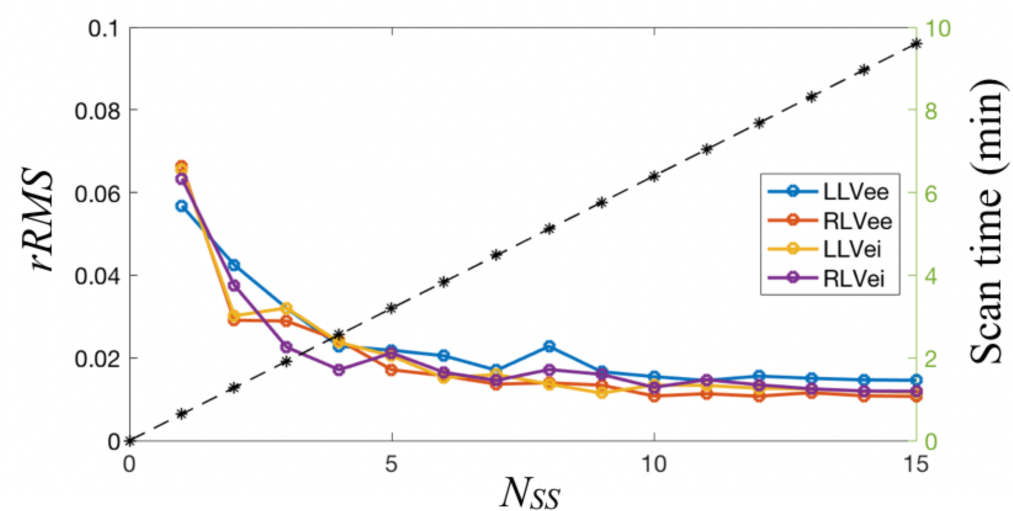
Illustration of relative root mean squared (*rRMS*) error of model fit for the 4 static lung volumes RLVei, LLVei, RLVee, and LLVee as a function of the parameters *N_SS_*, *MSL*, and *Int* in the modeling stage. *N_SS_* = Number of Slices Selected. *MSL* = Method of Selecting Locations. *Int* = Interpolation method. RLVei = Right Lung Volume at end-inspiration. RLVee = Right Lung Volume at end-expiration. LLVei = Left Lung Volume at end-inspiration. LLVee = Left Lung Volume at end-expiration.

**Figure S2.**
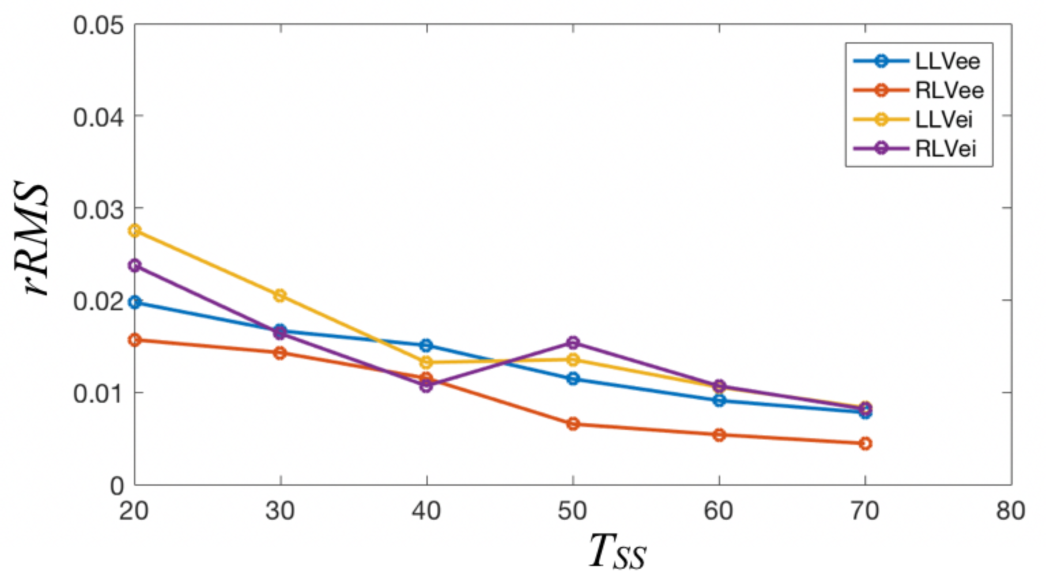
Illustration of relative root mean squared (*rRMS*) error of model fit for the 4 volumes RLVei, LLVei, RLVee, and LLVee and as a function of the parameter *T_SS_* in the modeling stage. RLVei = Right Lung Volume at end-inspiration. RLVee = Right Lung Volume at end-expiration. LLVei = Left Lung Volume at end-inspiration. LLVee = Left Lung Volume at end-expiration. *T_SS_* = Number of Time samples in Sparse Scan.

